# In Vivo Cortical Microstructure: A Proxy for Tauopathy and Cognitive impairment in the Elderly with and without MCI/Dementia

**DOI:** 10.1101/2021.03.26.21254351

**Authors:** John A. E. Anderson, Christin Schifani, Arash Nazeri, Aristotle N. Voineskos, for the Alzheimer’s Disease Neuroimaging Initiative

## Abstract

Aggregation of hyperphosphorylated tau protein is currently one of the most reliable indicators of Alzheimer’s pathology and cognitive impairment in older adults. However, it would be useful to have a non-invasive, accessible proxy measure that does not rely on Positron Emission Tomography (PET). We used data from multi-shell diffusion-weighted imaging (DWI) to assess indices from the Neurite Orientation Dispersion and Density Imaging (NODDI) model to determine possible proxies for tau and relationship with cognitive impairment. After controlling for age, sex, and the time difference between the scan acquisitions (DWI vs. PET), we used multiple factor analysis (MFA) to assess the fit between NODDI indices (orientation dispersion [ODI], neurite density [NDI], and free-water [fISO]), cortical thickness, and tau binding (via PET). We used data from 80 participants from the ADNI-3 sample who had a multi-shell DWI and an [^18^F]AV-1451 (tau) PET scan. Of these 80, 49 individuals were considered cognitively normal older adults (age ~74 years), 26 individuals had a diagnosis of mild cognitive impairment (age ~75 years), and five individuals had Alzheimer’s dementia (age ~78 years). fISO and tau shared a large amount of spatial overlap, and both strongly correlated with the first MFA dimension. Macrostructural features (such as cortical thickness and subcortical volume) introduced in a follow-up analysis were less related to this first MFA dimension than fISO and eight percent less than tau. Subsequent mediation analyses demonstrated that fISO mediated the relationship between cortical thickness and tau, explaining all of the variance. Microstructural features derived from advanced DWI acquisitions such as fISO may be useful proxies for tau. Cortical fISO, rather than cortical thickness, may represent the impact of tau on the brain (and, by extension, cognition).

## Introduction

Tau protein, an essential component of microtubule architecture in neurons and glia, can misfold due to hyperphosphorylation and aggregate into protein clumps and neurofibrillary tangles (NFT)^1^. These misfolded tau aggregates are the hallmarks of tauopathies, including Alzheimer’s disease (AD). The presence of tau aggregates in the brain in AD patients signals a transition from amyloidosis, characterized by amyloid fibrils building up in the brain tissue leading to cortical thickening and reduced free-water (i.e., fISO), to cortical thinning and cognitive impairment as NFTs accumulate^2^. Hyperphosphorylation of tau is initiated and exacerbated by oxidative stress as an immune response to acute or low-grade chronic inflammation, conceivably a secondary response to increasing amyloid-beta (Aβ) burden^1^. The presence of tau accumulation in the temporal lobes is more strongly associated with cognitive decline than amyloid in the region, suggesting it is a more direct measure of the clinical progression of AD^3^. It has been suggested that misfolded tau proteins can be exchanged at a cellular level and rapidly spread from subcortical to cortical regions by scaffolding highly connected regions such as the default mode network, moving to the prefrontal cortex later in AD progression^4–6^.

Imaging tau pathology is possible *in vivo* in the human brain using positron emission tomography (PET) with several widely used radioligands designed to bind the tau complexes such as [^18^F]MK-6240, [^11^C]PBB3, and [^18^F]AV-1451 flortaucipir; ^7^. While the ability of PET to target specific protein binding sites makes it invaluable, its use is restricted by the time-limited availability of radioligands. For example, Fluorine-18 (F-18)^8 9^ has a halflife of ~110 minutes, while Carbon-11 (C-11) is even shorter with ~20 minutes ^8^, thus making PET imaging challenging without a nearby cyclotron. PET imaging is also much more expensive^10^ than, for instance, MRI, and therefore its use is mainly restricted to research centers rather than clinical settings. Consequently, there is considerable interest in developing a more widely available, non-invasive, easily affordable proxy measure for tau. A more rapidly, inexpensive, and widely deployable method would facilitate longitudinal monitoring of patient progress, intervention as early as alterations in brain structure become apparent, and tracking of high-risk populations (including siblings, those with behavioral symptoms, and those with a genetic risk) for signs of early brain disruption. Monitoring indicators of cortical tau accumulation at this scale would also allow for more rapid clinical turnaround and intervention at earlier, potentially more treatable, stages.

Recent advances in diffusion-weighted imaging (DWI) have led to the development of biophysically plausible models that allow for modeling complex brain microstructure (including within the cortex) with only a minimal extra need for data collection compared to general DWI sequences. Unlike diffusion tensor models, which measure water diffusion over an entire voxel, biophysical models estimate water diffusion on much smaller scales and infer sub-voxel microstructural properties. Among such models, neurite orientation dispersion and density imaging ^NODDI; 11^ models water diffusion in three separate microstructural compartments (intracellular, extracellular, and CSF), providing independent measures of: (i) *neurite density* by estimating the fraction of water restricted in the intracellular compartment (neurite density index [NDI], ranges from 0-1 where higher values reflect greater neurite density), (ii) *neurite organization* by estimating the branching or dispersion of neurites (orientation dispersion index [ODI], ranges from 0-1 where higher values indicate greater dispersion), and the fraction of free water [fISO], indexing the percentage of the volume in each voxel that fISO occupies. While NDI is suggested to indicate cortical myelin, ODI likely indicates dendritic arborization and is relatively more expressed than NDI in the cortex^12–14^.

The combination of amyloidosis and tau aggregates results in cortical extracellular matrix expansion and changes in diffusion properties^15–18^. In AD compared to healthy aging control, lower cortical ODI and NDI values have been shown in temporal and frontal lobes^19,20^. Reductions in cortical NDI were associated with impaired neuropsychological performance in AD patients^19–22^, and ^18^F-THK5351 was shown to negatively correlate with ODI and NDI in gray matter in ^11^C-PiB+ patients^23^. In line with the human *in vivo* imaging studies, rodent *in vivo* imaging has shown that tau-mouse models have lower cortical and hippocampal NDI values than wild type mice, and NODDI metrics tracked histopathology better than DTI-derived measures^21^.

Increases in fISO have been linked to the presence of AD in subcortical grey matter such as the hippocampus^24^ and cortically in temporal, posterior cingulate, medial prefrontal, and parietal regions^2^. FISO is thus well-positioned as a prospective marker for the presence of tau. Tau PET has also been shown to be the best currently available in vivo biomarker of AD pathology, and its presence is much more tightly linked to cognitive decline^25–27^. While studies have independently identified alterations in NODDI metrics and increases in tau binding in AD, the multivariate relationship between microstructural NODDI indices, cognitive outcomes, and tau deposition in gray matter has not yet been explored. In the present study, using a multivariate approach known as multiple factor analysis (MFA), we examine how microstructural indices of brain health covary with tau deposition and cognition, hypothesizing that NODDI would reveal an MRI proxy for tau with sensitivity to the presence of MCI. The main advantage of MFA is to assess the multivariate relationships between brain and behavior indices rather than examining each serially. Thus we are taking a systems rather than reductionist level approach to the aging brain. Specifically, we expect to find a component describing a covariance pattern showing worse cognitive performance is associated with higher tau and fISO and lower NDI and ODI. Furthermore, by using a multivariate approach, we can also disambiguate the shared and unique contributions of different MRI measures (e.g., fISO vs. cortical thickness) to variance in tau and cognitive performance.

## Materials and methods

### Participants

Data used in this article were obtained from the ADNI database (adni.loni.usc.edu). Within the ADNI database, 640 individuals had an [^18^F]AV-1451 (tau) PET scan, and 103 individuals had a multi-shell DWI scan, while 94 individuals had both. After removing duplicates and poor data, 80 individuals had both high-quality data for one tau and one multi-shell DWI scan and also had neuropsychological assessments (downloaded 05/04/2019). This sample of 80 individuals with all three measures (tau, multi-shell DWI, and neuropsychological measures) was retained for analysis (see Table 1; note neuroimaging variables are raw scores, not residuals). In this sample, 49 individuals were cognitively normal (CN, mean age = 73.8, range 62:90 years), 26 had a diagnosis of MCI (mean age = 75, range 60:91), and 5 had an AD diagnosis (mean age = 78, range 69:83).

**Table 1:** Sample descriptive statistics. Means and standard deviations (in parentheses)

|  | CN (N=49) | MCI (N=26) | AD (N=5) |
| --- | --- | --- | --- |
| **Sex (Number of females)** | 34 (69.4%) | 6 (23.1%) | 3 (60%) |
| **MB age** | 73.8 (7.66) | 75.3 (6.8) | 77.8 (5.86) |
| **ADNI Executive Function Score** | 1.07 (0.85) | 0.530 (0.80) | -1.16 (0.84) |
| **ADNI Memory Score** | 0.85 (0.50) | 0.43 (0.66) | -1.15 (0.53) |
| **Date Difference (in days)** | -164 (192) | -103 (338) | -172 (227) |
| **Tau** | 239 (2.72) | 238 (4.41) | 240 (2.68) |
| **Cortical Thickness** | 619 (69.9) | 619 (59.1) | 561 (70.8) |
| **fISO** | 0.12 (0.03) | 0.14 (0.03) | 0.19 (0.04) |
| **ODI** | 0.44 (0.01) | 0.44 (0.02) | 0.42 (0.01) |
| **NDI** | 0.48 (0.02) | 0.48 (0.02) | 0.48 (0.02) |

### Cognitive measures

We elected to use the factor scores for memory and executive function supplied with the ADNI dataset^28–30^ rather than raw neuropsychological scores. The memory factor score incorporates the Rey Auditory Verbal Learning Test (RAVLT), the Alzheimer’s Disease Assessment Scale-Cognitive Subscale (ADAS-Cog), the Logical Memory Test, and the Mini-Mental Status Examination (MMSE). In contrast, the executive function factor score includes category fluency (animals and vegetables), the Trail Making Test A/B, Digit Span, Digit Symbol, and the Clock Drawing Test. These scores are driven by the inclusion of measures based on theory, have been well documented and validated, and are widely reported in the ADNI literature. Using the composite scores, we also reduce the complexity of the interpretation (in the multivariate analysis) and the need for multiple comparison corrections (in our univariate analyses). Finally, factor scores are representative of the global effects on memory and executive function, which are more pertinent for our analysis than scores on individual neuropsychological measures.

### Tau PET acquisition and preprocessing

We downloaded fully-processed PET data from the ADNI database. PET scans were acquired on a Siemens Biograph TruePoint PET/CT scanner beginning at 75 min post-injection of 370 MBq (10.0 mCi) ± 10% [^18^F]AV-1451, for 30 min (6 × 5 min frames) in list mode as described before^31^. Reconstructions were performed per ADNI-3 protocol, as defined by the Laboratory of Neuro Imaging (LONI).

Processing was done by UC Berkeley, as previously described^31–33^. In brief, [^18^F]AV-1451 data were downloaded from LONI in the most fully pre-processed format, realigned, and the mean of all frames was used to co-register [^18^F]AV-1451 data to each participant’s structural MRI. [^18^F]AV-1451 standardized uptake value (SUV) images were created based on mean uptake over 80–100 min post-injection normalized to uptake in a gray matter-masked cerebellum reference region (inferior cerebellar gray matter) to create voxelwise SUV ratio (SUVR) images in each participant’s MRI native space. SUVR images were smoothed to a standard resolution of 8 mm^3^ (Joshi et al., 2009). [^18^F]AV-1451 SUVR images were corrected for partial volume (PV) effects using the Geometric Transfer Matrix approach ^34^ for PVC based on FreeSurfer-derived ROIs ^33^. The PVC goals were to correct for choroid plexus, and basal ganglia signal bleeding into neighboring regions (such as the hippocampus) and account for PV effects due to atrophy. Additionally, skull, tissue, and CSF segments were used from SPM tissue probability masks to correct for high signal in extra-cortical regions. Finally, ROI-specific PV-corrected SUVR values were re-normalized by PV-corrected inferior cerebellar gray matter^31^.

### Multi-shell diffusion imaging preprocessing

Raw multi-shell and T1-weighted images acquired in the same session were downloaded from the ADNI database. All scans were acquired as part of the ADNI-3 protocol from the same site using a 3-Tesla Siemens MAGNETOM Prisma scanner. The structural scan was acquired with a 13 min 52 s MPRAGE sequence with 1.0 mm^3^ resolution (TR = 2300 ms, TE = 2.98 ms, inversion time = 900 ms, flip angle = 9 degrees). The multi-shell DWI images were acquired with a single-shot, multiband, multi-shell acquisition (TR = 3400 ms, TE = 71 ms, flip angle = 90 degrees, voxel size = 2.0 mm^3^ isotropic, FOV = 116 mm, acquisition time = 14 minutes 31 seconds, acceleration factor = 3, phase-encoding direction = anterior to posterior) with 2 DWI shells b=1000 s/mm2 (48 directions) and b = 2000 s/mm2 (60 directions) and additional 13 b=0 s/mm2 volumes interspersed throughout the acquisition. Cortical reconstruction and volumetric segmentation of MPRAGE images were performed with Freesurfer version 6 (http://surfer.nmr.mgh.harvard.edu/) as described in prior publications ^e.g., 35^. Downloaded multi-shell DWI scans were denoised using Marchenko–Pastur Principal Component Analysis (MP-PCA) denoising^36,37^ using the dwidenoise command in MRtrix v. 3.0^36,38^ then preprocessed in a single step using eddy^39^ (version 5.0.11) with slice-wise outlier replacement^40^, simultaneously correcting eddy current-induced distortions and subject movement, thus allowing for correction of higher b-values, inherently having a lower signal-to-noise ratio. Phase-encoding distortion was corrected post-hoc by non-linearly registering each person’s first b0 image to their structural data constrained to the A-P direction (using the bdp pipeline in Brainsuite v.18a) ^41^.

The NODDI model was then fit with the Microstructure Diffusion Toolbox (https://github.com/robbert-harms/MDT) to each participant’s preprocessed DWI data^11,42^, producing NODDI parameter maps. The NODDI indices were brought into surface space e.g., aligned to the 32K midthickness FreeSurfer outputs; ^43^, and average midthickness values were extracted for each index per each ROI of the Desikan-Killiany-Tourville (DKT) atlas^44^.

### Analysis

#### Multiple Factor Analysis

First, age, sex, and the time between tau PET and multi-shell scans (see Table 1 for average time between scans) were regressed out of each variable of interest, and the residuals were saved as inputs for the next step. A multiple factor analysis (MFA) was then used to examine the multi-table (e.g., each imaging modality, cognition, diagnosis, etc.) relationship between each of the NODDI indices (ODI, NDI, fISO) and tau. Cognition (memory and executive function) and diagnostic group (CN, MCI, or AD) were treated as supplementary variables that did not determine the factor solution. MFA is an extension of principal components analysis or factor analysis. MFA can be considered a *weighted PCA* where each set of variables undergoes a PCA analysis. Contributing variables are weighted by dividing them by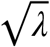, and a superordinate PCA estimates the relationship between sets of variables. MFA extends traditional PCA from a two-table to a multi-table solution, allowing users to analyze interactions of sets of variables and preserve the grouped structure (e.g., demographics, cognitive variables, brain imaging variables), and crucially, the balance between sets of variables. Considering brain imaging, which could conceivably have hundreds or thousands of variables, MFA balances each of the tables’ contribution to the analysis such that one factor does not dominate the study through the sheer number of observations.

The number of components to retain was determined by examining eigenvalues larger than one (i.e., a scree plot) and testing whether the observed eigenvalue for the dimension of interest was different from chance using 500 random permutations. To determine reliable contributions to each dimension of the MFA solution, we performed a bootstrapping procedure (1000 repetitions), from which we calculated a bootstrap ratio (e.g., a ratio of the MFA contribution to the estimated standard error from bootstrapping). Therefore, higher bootstrap ratios contribute more to the latent dimension having a higher weight and smaller variance ^45,46^. Bootstrap ratios can be interpreted similarly to a z-score, and we selected a threshold of 2 to indicate reliable values^47^. The strengths of multivariate approaches to wide data such as in neuroimaging or genetics are their ability to increase power by testing sets of covarying features rather than individual tests at each ROI. Since multivariate approaches analyze covariance structures over regions and modalities, they also capitalize on the fact that the brain is organized as a network. Using multivariate methods changes the focus from which region or regions vary per data type across individuals to what sets of regions and data types covary across individuals; thus, multivariate techniques allow various data types and regions to inform one another, producing a more cohesive result.

As cortical thickness is often considered a macrostructural indicator of cognitive decline and dementia, we repeated the above analysis, including FreeSurfer extracted cortical thickness and subcortical volume estimates to examine how these indices covaried with NODDI indices and tau.

#### Mediation Analysis

While grey matter ODI and NDI are, in theory, not affected by partial volume effects from CSF, fISO is inherently related to partial volume effects. In the presence of brain atrophy, such as in AD, gray matter fISO could conceivably be higher simply because there is more CSF partial volume with macrostructural atrophy. Therefore, it is important to estimate the contribution of fISO beyond these macrostructural changes. To do this, we extracted summary principal component scores for tau, fISO, and cortical thickness/subcortical volume for each person across all brain regions. We then used the *psych* package in R^48^ to fit a mediation analysis using fISO as the predictor, tau as the dependent variable, and cortical thickness/ subcortical volume as the mediator. If macrostructural changes drive the increase in fISO, then including macrostructural features in a model should eliminate fISO’s relationship with tau.

#### Partial correlations

While the MFA allows us to examine a global portrait of how tau interacts with NODDI indices and macrostructural measures, it is still useful to explore the univariate relationships using partial correlations (e.g., tau and ODI or NDI or fISO). As univariate approaches only consider local covariance within regions of interest (ROIs) and between the modalities of interest without considering other indices, we might expect similar but not identical results to MFA. Data were analyzed using R version 3.6.2 (2019-12-12), “Dark and Stormy Night”^49^ using the FactoMineR and factoExtra packages for MFA and HCPC^50,51^, and brain data were visualized with ggseg^52^.

Partial correlations between tau and NODDI indices were calculated, controlling for age, sex, and the time difference (in days) between the tau PET and multi-shell scans (see Table 1 for average time between scans). Correlations were Bonferroni-corrected (*p* <0.05_corrected_) for multiple comparisons (80 ROIs), and bootstrapped (1000 repetitions) to estimate 95% confidence intervals. Lack of overlap of the confidence intervals can thus be reasonably interpreted as a reliable difference between correlations. Partial correlations between tau or NODDI indices with summary factor scores of cognition (executive function and memory) were also calculated as described above.

#### Data availability

The data that support the findings of this study are openly available in the ADNI repository at http://adni.loni.usc.edu/.

## Results^1^

### Multiple Factor Analysis I: How do tau and NODDI indices Covary?

Multiple factor analysis yielded two significant dimensions with eigenvalues larger than one (both *p’s* < 0.001; λ_1_ = 1.77, λ_1_ = 1.53) that explained 16.3% and 14.1% of the variance (Figure 1; see also Supplementary Tables 1 and 2). See Supplementary Table 3 for ROI names.

**Figure 1.**
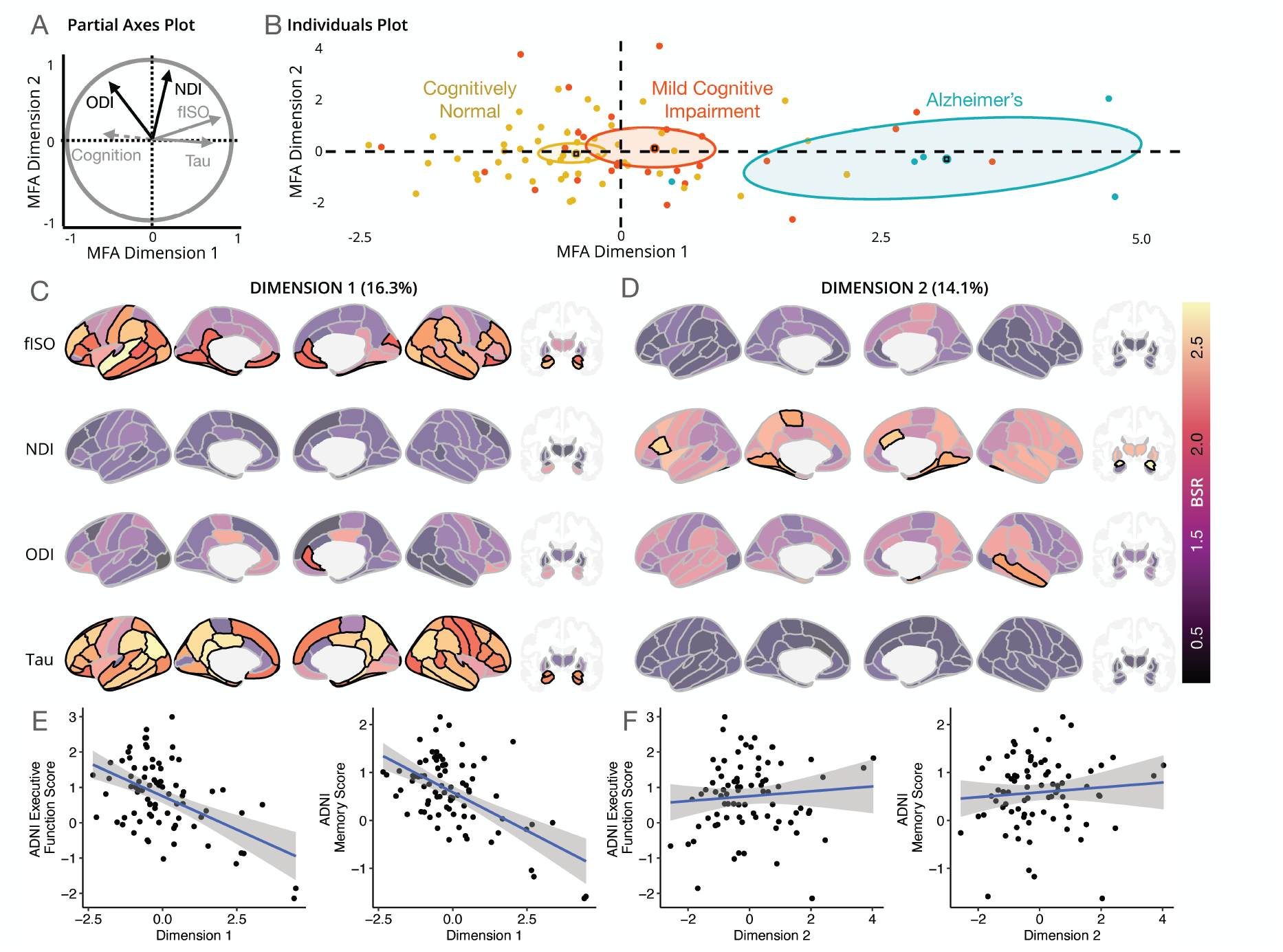
Multiple factor analysis (MFA) of NODDI indices (fISO, NDI, ODI), and tau, with cognition (executive function and memory) and diagnosis (cognitively normal, mild cognitive impairment, Alzheimer’s Disease) as supplementary variables. Panel A) shows partial axes for each variables’ first dimension plotted against the first two dimensions of the MFA; lines and text in black highlight variables that align more with the second dimension. Cognition is shown with a broken line to indicate it did not contribute to the MFA solution. Panel B) shows individuals plotted along the first two dimensions supplemented by colors indicating group membership. 95% confidence ellipses surround group centroids, and lack of overlap suggests a reliable group difference. Panels C)and D) show bootstrap ratio (BSR) maps of brain regions by modality contributions to the first and second dimensions. Solid colors with black outlines indicate reliable associations (e.g., BSR > 2), other regions are shown to provide context. Panels E) and show correlations between the MFA dimensions and the composite neuropsychological measures for the first and second dimensions. ADNI = Alzheimer’s Disease Neuroimaging Initiative; fISO = free-water; NDI = neurite density index; ODI = orientation dispersion index.

**Figure 2.**
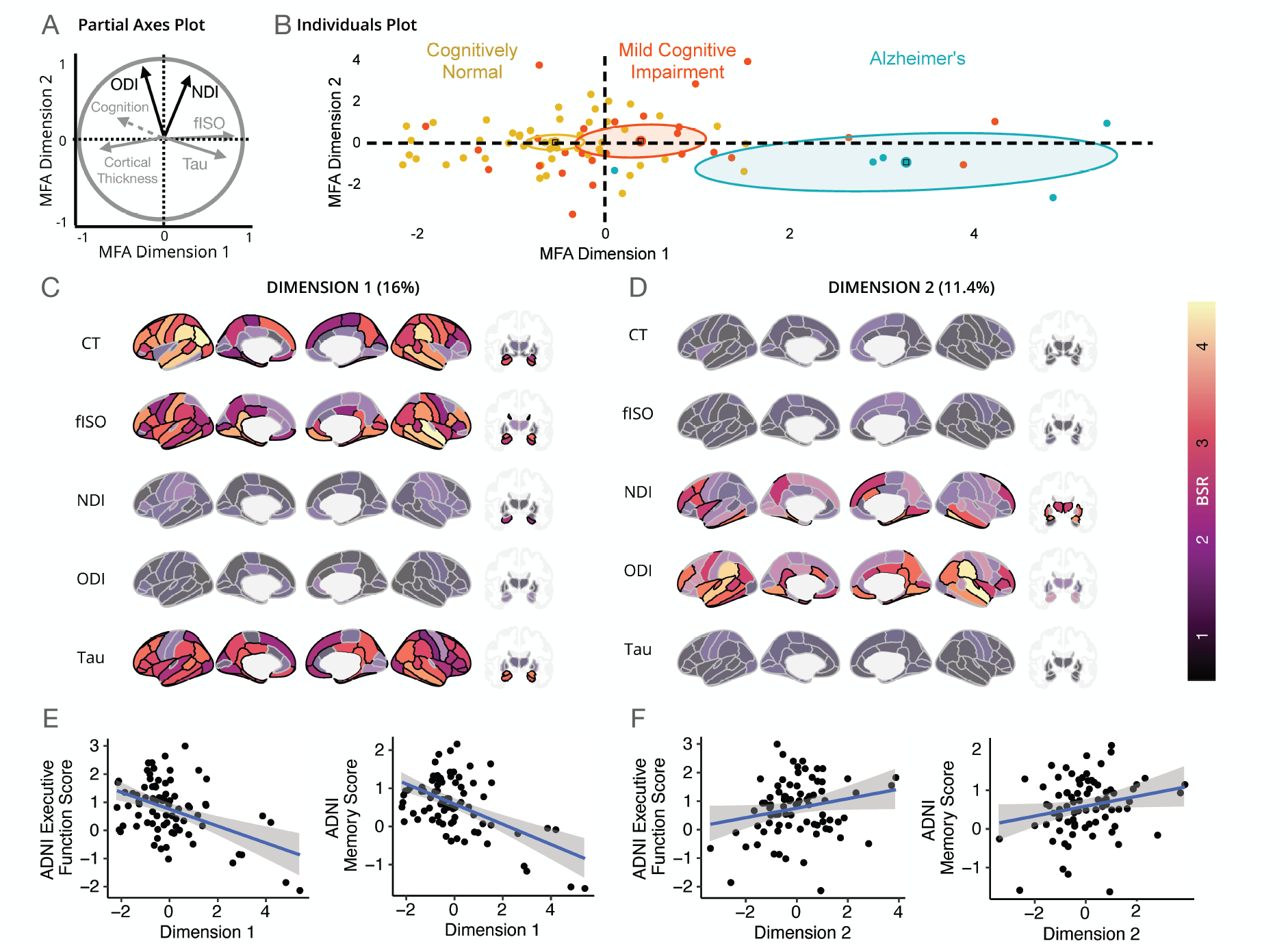
Multiple factor analysis (MFA) of NODDI indices (fISO, NDI, ODI), and tau and cortical thickness (CT), with cognition (executive function and memory) and diagnosis (cognitively normal, mild cognitive impairment, Alzheimer’s Disease) as supplementary variables. Panel A) shows partial axes for each variables’ first dimension plotted against the first two dimensions of the MFA; lines and text in black highlight variables that align more with the second dimension. Cognition is shown with a broken line to indicate it did not contribute to the MFA solution. Panel B) shows individuals plotted along the first two dimensions supplemented by colors indicating group membership. 95% confidence ellipses surround group centroids, and lack of overlap suggests a reliable group difference. Panels C)and D) show bootstrap ratio (BSR) maps of brain regions by modality contributions to the first and second dimensions. Solid colors with black outlines indicate reliable associations (e.g., BSR > 2), other regions are shown to provide context. Panels E) and F) show correlations between the MFA dimensions and the composite neuropsychological measures for the first and second dimensions. ADNI = Alzheimer’s Disease Neuroimaging Initiative; fISO = free-water; NDI = neurite density index; ODI = orientation dispersion index.

Average tau and fISO strongly correlated with the MFA’s first dimension (*r*_*tau*_ = 0.53, *r*_*fISO*_ = 0.50), while ODI and NDI did not (*r*_*ODI*_ = − 0.26, *r*_*NDI*_ = 0.13); see partial axes plot in Figure 1A. Bootstrap ratio values in Figure 1C show regions that contributed to the first dimension. Tau ROIs reliably contributing to the first dimension included bilateral hippocampus and amygdala, temporal and parietal regions, posterior cingulate, and bilateral dorsolateral prefrontal cortex. Spatial patterns of fISO covariance were similar to tau, though more anatomically constrained. For example, fISO covaries with the first dimension in the isthmus of the cingulate gyrus, while this covariance pattern extends into the posterior cingulate cortex for tau.

Conversely, higher fISO values are expressed in the lateral and medial orbitofrontal cortex and rostral anterior cingulate, where tau contributes less variance. Notably, the rostral anterior cingulate cortex is also a region with lower ODI values. The MFA’s first dimension also negatively correlated with the two auxiliary cognitive variables (executive function and memory factors; Figure 1E; *r_Executive Function_* = − 0.51, *r_Memory_* = − 0.58), suggesting that higher values on the first dimension generally predicted worse cognition (see Supplemental Figure 1 for the parameter-specific PCAs contributing to the MFA).

The second MFA dimension correlated most robustly with NDI and ODI and not with tau or fISO. NDI regions contributing reliably to this second dimension included the bilateral fusiform gyrus, bilateral lingual gyrus, and bilateral parahippocampal gyrus(see Figure 1D). Other reliable NDI regions were left amygdala, left pars opercularis, right caudal anterior cingulate, left paracentral gyrus, and right putamen.

ODI contributed reliably to the right entorhinal and middle temporal cortices. However, bootstrap ratios associated with this dimension were sparse and were not related to diagnostic severity or cognition (Figure 1F; but see Supplemental Figure 2 for hierarchical clustering on principal components containing a subgroup whose levels of dimension 2 predicted cognition).

### Multiple Factor Analysis II: How do macrostructural features covary with NODDI and tau?

We next extended the initial multiple factor analysis to include cortical thickness and subcortical volume estimates (see Supplemental Figure 1). This analysis again yielded two significant dimensions with eigenvalues larger than one (both *p’s* < 0.001; λ_1_ = 2.25, λ_1_ = 1.60) that explained 16% and 11.4% of the variance. Results from this analysis were remarkably similar to the MFA results without cortical thickness. Notably, cortical thickness and subcortical volume loaded negatively onto the MFA’s first dimension capturing general atrophy (i.e., in opposition to fISO and tau, *r*_*CT*_ = − 0.41, *r*_*fISO*_ = 0.50, *r_tau_* = 0.52). As in the initial MFA, the first MFA dimension correlated with the two auxiliary cognitive variables (*r_Executive Function_* = − 0.45, *r_Memory_* = − 0.54).

Including the macrostructural features improved the second dimensions’ correlations with the supplementary cognitive variables (*r_Executive Function_* = 0.21, *r_Memory_* = 0.22) compared with the initial MFA (*r_Executive Function_* = 0.08, *r_Memory_* = 0.08). Higher scores on this second MFA dimension suggests that higher memory and executive function scores may reflect higher levels of cortical NDI in the bilateral prefrontal cortex, inferior medial temporal lobe, and higher subcortical values of NDI in the bilateral thalamus, putamen, and amygdala, and higher cortical ODI scores in temporal, posterior cingulate, and frontal regions.

### Mediation Analysis: does fISO predict tau after controlling for cortical thickness/ subcortical volume?

Figure 3A shows that introducing cortical thickness as a mediator does *not* significantly impact the significant relationship between fISO and tau, and cortical thickness/subcortical volume mediates the effect of fISO on tau by only 1.12%. Conversely, Figure 3B shows that the path from cortical thickness through fISO to tau mediates the relationship between cortical thickness and tau by 29.9%.

**Figure 3.**
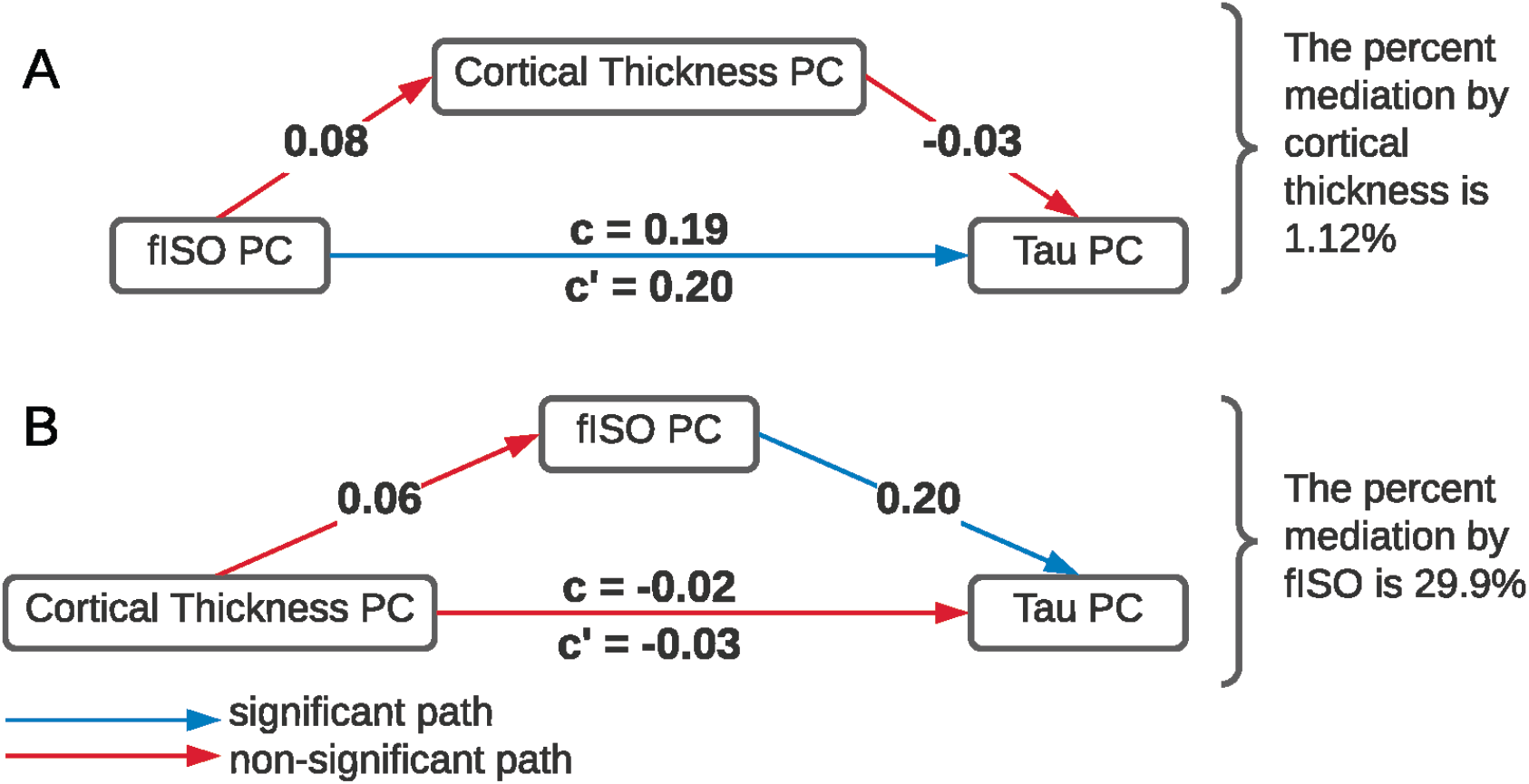
Mediation analyses between principal component (PC) scores for fISO (free-water), cortical thickness (CT), and tau. Panel A) shows the effect of controlling for cortical thickness when predicting tau from fISO. Panel B) shows the effect of controlling for fISO when predicting tau from cortical thickness.

### Partial Correlations by Modality: Considering each modality on its own

Univariate partial correlations between each NODDI parameter and tau are shown in Figure 4A. In those ROIs where NODDI parameter/tau relationships were significant after within-modality multiple comparison correction, we also show univariate relationships with ADNI factor scores for memory and executive function(see Figure 4B).

**Figure 4.**
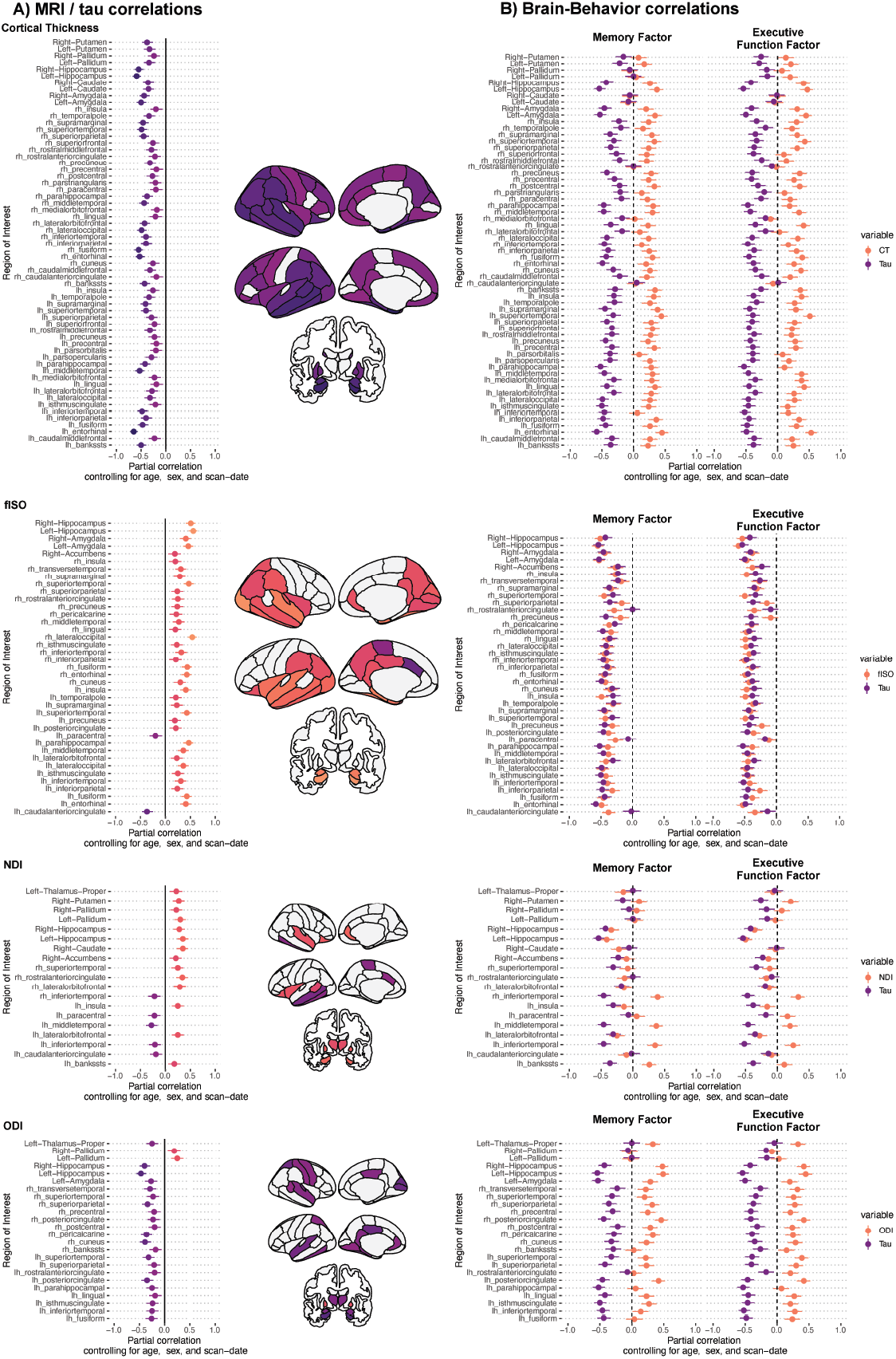
Surfaces and point-range plots of partial correlations between MRI indices and tau. C) Partial correlations between each MRI parameter or tau and ADNI memory or executive function factor scores. All error bars are bootstrapped 95% confidence intervals, and reliable differences can be inferred by lack of overlap. ADNI = Alzheimer’s Disease Neuroimaging Initiative; CT = cortical thickness; fISO = free-water; lh = left hemisphere; NDI = neurite density index; ODI = orientation dispersion index; rh = right hemisphere.

We observed that fISO and tau positively correlated in the bilateral hippocampus and amygdala, bilateral temporal and parietal regions, and bilateral precuneus and posterior cingulate. Two regions (left paracentral lobule and left caudal anterior cingulate) countered the global trend and expressed negative associations. In regions where fISO and tau were positively correlated, each had negative associations with memory and executive function. Memory was not reliably predicted by tau in three regions, left paracentral lobule, right rostral anterior cingulate, and left caudal anterior cingulate (i.e., the 95% bootstrapped CI overlapped with zero), and fISO more reliably predicted memory than tau in the latter two regions. Executive function was more reliably predicted by fISO in the right rostral anterior cingulate, while tau more reliably predicted executive function in the right precuneus.

Cortical thickness and subcortical volume was globally negatively associated with tau and did not show regional variation. Higher levels of cortical thickness were associated with higher memory and executive function scores.

ODI was negatively associated with tau in the bilateral left hippocampus, amygdala, thalamus, superior temporal lobes, and right pre and postcentral gyri. Tau and ODI also negatively correlated in regions including the left posterior cingulate and rostral anterior cingulate. Conversely, positive correlations between tau and ODI were observed in bilateral pallidum. For each of these regions, the global pattern was for higher ODI values to predict better executive function and memory task performance, while the converse was true for tau. In contrast to fISO, most of the correlations between ODI and cognition were reliably different from tau and cognition (seen from the lack of overlap of the 95% bootstrapped confidence intervals on the correlations). An exception is the bilateral pallidum, where the tau and ODI did not differ in their relationships with either memory or executive function.

Finally, NDI produced a more complex pattern of results. Higher NDI values were also associated with higher tau values in subcortical regions, including the bilateral thalamus, hippocampus, and pallidum. However, there were also negative associations in bilateral inferior temporal, left middle temporal, and left caudal anterior cingulate and paracentral lobes. How tau and NDI predicted behavior also varied regionally, with higher values predicting worse memory and executive function in the bilateral hippocampus, opposite results observable in specific cortical regions, including the bilateral inferior temporal cortex.

## Discussion

In the present study, we used a sample with a range of cognitive ability from the ADNI dataset and a multivariate approach to examine the relationship between tau and indices derived from NODDI, a biophysically plausible model based on DWI data. Our goal was to explore the potential of NODDI-metrics, especially fISO, as a biomarker for the disease progression of cognitive impairment with similar specificity to tau. We showed that the first dimension of an MFA captured a robust positive relationship between fISO and tau, strongly associated with worse cognition and diagnosis. Regions contributing to this pattern were those commonly implicated in cognitive decline and the progression of AD. They included the bilateral hippocampus, temporal lobes, and posterior cingulate, agreeing with models suggesting neuropathology can spread through highly connected regions of the default mode network. A second MFA dimension captured much of NDI and ODI variation but was not associated with cognition. A second MFA incorporated brain macrostructure, i.e., cortical thickness, strongly associated with tau and fISO. While a mediation model showed the association between tau and cortical thickness was eliminated by including fISO, including cortical thickness in the MFA increased the explanatory power of NDI and ODI on the second dimension and boosted their association with behavior, suggesting that cortical thickness is a suppressor variable. We conclude that fISO may be a suitable, minimally invasive proxy measure for tau pathology in dementia.

The role of the second MFA dimension capturing higher neurite density and orientation dispersion values, which have previously been linked to greater myelination and dendritic arborization, respectively, within the cortex, is potentially driven by macrostructural features, clarified by our second MFA (see Figure 2). Regions associated with the second MFA dimension initially included bilateral fusiform gyrus, temporal lobes, and the anterior cingulate. Including cortical thickness in the MFA expanded and clarified this pattern to bilateral temporal lobes, posterior cingulate bilateral frontal regions, and bilateral subcortical areas (thalamus, putamen, and amygdala for NDI). Thus including macrostructural features allowed the second MFA dimension to capture variance in regions disproportionately vulnerable to age-related cognitive decline. In the second MFA, which included macrostructural features (i.e., cortical thickness and subcortical volume), ODI and NDI still loaded onto the second dimension; however, now higher values on this dimension were associated with more robust cognition. This was further supported by our cluster analysis of the first MFA (see supplementary Figure 2). Two of the clusters separated along the second MFA dimension. Within the cluster of individuals with lower values of the second MFA components representing lower dendritic arborization and neurite density values, those with higher scores on this dimension had higher executive function performance. This suggests that cognitive differences in early-stage cognitive decline and healthy aging may instead be captured by measures of neurite density (i.e., myelination) and orientation dispersion (i.e., dendritic arborization) than fISO and tau.

To further decompose each NODDI index’s specific relationship with tau, we conducted partial correlations that largely replicated the above findings from the MFA, e.g., reliable positive correlations between tau and fISO in temporal, parietal, and hippocampal regions - (though the relationship between these variables in the dorsolateral prefrontal areas was not apparent). Cortical thickness and ODI showed the expected negative relationship with tau, suggesting that cortical dendritic branching and tissue density decreased as tau presence increased. Univariate comparisons showed cortical thickness was related to tau across more regions than any other measure. In contrast with tau, both higher cortical thickness and ODI values were also associated with better memory and attention performance. Finally, NDI produced a more complex set of results regarding the relationship between tau and behavior.

Our findings underscore the value of multivariate analyses compared to univariate analyses. A multivariate approach’s strength is that covarying data patterns can produce much more realistic, albeit complex, portraits of parameter relationships that may more accurately capture in vivo interactions. While we observed significant univariate associations of tau to NDI and ODI, these indices’ contribution was minimized when considered in a multivariate context. This, however, may not be the case for healthy younger adults or when only considering a sample of healthy aging individuals where pathology is less salient, though it remains unclear since multivariate multimodal imaging is not yet routinely applied to NODDI outputs(though see^53^) and future studies are warranted.

When considering whether fISO might be a suitable proxy measure for tau, we have established that both indices share a remarkable amount of spatial overlap via both the univariate and multivariate analyses. However, cortical thickness (and subcortical volume) also overlaps considerably with tau and fISO, which is why it is often used as a reliable macrostructural indicator of dementia and disease progression. Using mediation analysis, we could clearly show that cortical thickness and subcortical volume cannot explain fISO’s relationship with tau, and thus suggests that microstructural fISO is a stronger proxy for tau propagation than cortical thickness and subcortical volume. This is also reflected in our second MFA analysis (Figure 2). The first MFA component accounts for 71% of the variance in fISO and 72% of the variance in tau but only 64% of the variance in cortical thickness/subcortical volume. This suggests that there is only one percent separation between fISO and tau while there is an eight percent separation between tau and macrostructural features.

Within the cortex, both ODI and NDI were reduced in early AD pathology compared to healthy aging controls within a set of regions known to be especially vulnerable to AD pathology (such as entorhinal cortex, inferior temporal gyrus, middle temporal gyrus, fusiform gyrus, and precuneus)^19^ which was interpreted as consistent with a decrease in dendritic arborization and loss of dendritic spines and synapses. Wide-spread reductions in synaptic density (estimated by state-of-the-art in vivo PET imaging targeting the presynaptic vesicular protein 2A) have been indeed reported recently, including medial temporal and neocortical brain regions in early AD compared to healthy control participants ^54,55^ and they were found to be more extensive than decreases in gray matter volume ^54^. In line with that, Parker et al. reported ODI and NDI reductions were further found to be more strongly associated with diagnosis of AD compared to controls than cortical thickness a finding echoed by ^20^. We also show that increases in tau are broadly associated with decreases in ODI and NDI and increases in fISO. Our findings are supported by recent postmortem PET evidence showing a negative relationship between tau and synaptic density PET and a positive relationship between tau and neuroinflammation-related PET in the middle frontal gyrus (Brodmann area 46) of AD patient tissue ^56^. While tau aggregation reflects AD’s direct etiology, fISO likely captures downstream neuroinflammatory processes, which are useful measures of cognitive decline in their own right and can supplement tau PET imaging.

A common approach to using multishell DWI data is to eliminate the fISO fraction and examine corrected fractional anisotropy and mean-diffusivity; however, the free-water fraction itself appears to be more closely associated with the presence of CSF phosphorylated-tau and Aβ42 in temporal, parietal, and medial prefrontal white matter (homologs of many of the gray matter regions we report) ^57^ than these corrected indices. Within the cortex, the fISO fraction is proposed to follow a biphasic trajectory along the course of healthy aging to MCI and AD^2^. It is hypothesized that early in the period of dementia (i.e., stage one), amyloidosis results in cortical thickening and a reduction in fISO due to neuroinflammation and increased presence of glia and extracellular matrix expansion, which affects diffusion properties ^15–18^, while later in the disease (i.e., stages two and beyond), abnormal tau aggregates lead to cortical thinning and accumulation of fISO as cells atrophy and barriers to diffusion fail. This work highlights the importance of examining longitudinal changes in potential disease markers as their interpretation may change based on the disease stage. While previous studies have demonstrated correlations between dementia and fISO and between CSF measures of tau and fISO in dementia, our study highlights the specificity of the relationship between tau and fISO in a spectrum from healthy aging to dementia by using complementary in vivo PET and DWI. Notably, we demonstrate that fISO mirrors tau’s distribution in the hippocampus, temporal lobes, parietal lobes, and medial prefrontal cortices - all regions known to be especially vulnerable to cognitive decline and dementia progression^5,6^.

### Limitations and potential solution

Our study sample size was limited by the availability of multishell scans with concurrent [^18^F]AV1451 (tau) PET scans. In time, the ADNI dataset will accrue more scans, and we suggest this descriptive model be revisited and validated. We also acknowledge that relationships between tau and fISO and other neurite integrity measures may not be linear. Indeed, it has been proposed that there may be nonlinearities affecting fISO and cortical thickness, particularly in early AD^2^. Addressing nonlinear changes with time requires longitudinal data, which is currently being collected by the ADNI consortium but does not yet contain enough (replicate) data to address this question.

## Conclusions

Using multivariate modeling, we found that fISO (or free-water) derived from the NODDI model is associated with in vivo tau estimated with PET imaging. We also demonstrate that fISO and tau align and collectively explain more variation in cognition and diagnostic status included as auxiliary variables in the analysis. In mediation, the primary MRI in vivo variable, cortical thickness, previously considered to be a proxy for tau, was no longer influential in the context of fISO. Therefore, we conclude that hyperphosphorylated tau deposition can be effectively indexed using noninvasive MRI via increases in fISO. If replicated, our findings may open up possibilities for larger, less expensive clinical trials, aiming to prevent or delay dementia using noninvasive MRI and fISO as a biological endpoint.

## Acknowledgments

The authors are grateful to Dr. Peter Zhukovsky and Jovanka Skocic for their thoughtful comments on an earlier version of this manuscript. Data collection and sharing for this project was funded by the Alzheimer’s Disease Neuroimaging Initiative (ADNI) (National Institutes of Health Grant U01 AG024904) and DOD ADNI (Department of Defense award number W81XWH-12-2-0012). ADNI is funded by the National Institute on Aging, the National Institute of Biomedical Imaging and Bioengineering, and through generous contributions from the following: AbbVie, Alzheimer’s Association; Alzheimer’s Drug Discovery Foundation; Araclon Biotech; BioClinica, Inc.; Biogen; Bristol-Myers Squibb Company; CereSpir, Inc.; Cogstate; Eisai Inc.; Elan Pharmaceuticals, Inc.; Eli Lilly and Company; EuroImmun; F. Hoffmann-La Roche Ltd and its affiliated company Genentech, Inc.; Fujirebio; GE Healthcare; IXICO Ltd.; Janssen Alzheimer Immunotherapy Research & Development, LLC.; Johnson & Johnson Pharmaceutical Research & Development LLC.; Lumosity; Lundbeck; Merck & Co., Inc.; Meso Scale Diagnostics, LLC.; NeuroRx Research; Neurotrack Technologies; Novartis Pharmaceuticals Corporation; Pfizer Inc.; Piramal Imaging; Servier; Takeda Pharmaceutical Company; and Transition Therapeutics. The Canadian Institutes of Health Research is providing funds to support ADNI clinical sites in Canada. Private sector contributions are facilitated by the Foundation for the National Institutes of Health (www.fnih.org). The grantee organization is the Northern California Institute for Research and Education, and the study is coordinated by the Alzheimer’s Therapeutic Research Institute at the University of Southern California. ADNI data are disseminated by the Laboratory for Neuro Imaging at the University of Southern California.

## Supplementary Material

### Supplemental multiple factor analysis (MFA) information for the first MFA analysis (without cortical thickness)

In addition to the findings presented in the main part of the manuscript, we also present a table of R-vector (RV) coefficients showing the multivariate correlations between variables contributing to the MFA (across dimensions) and auxiliary variables (see Supplementary Table 1).

**Supplementary Table 1.**

|  | Diagnostic Status | Cognitive Scores | Tau | fISO | ODI | NDI | MFA |
| --- | --- | --- | --- | --- | --- | --- | --- |
| Diagnostic Status | 1 |  |  |  |  |  |  |
| Cognitive Scores | 0.31 | 1 |  |  |  |  |  |
| Tau | 0.20 | 0.25 | 1 |  |  |  |  |
| fISO | 0.24 | 0.23 | 0.21 | 1 |  |  |  |
| ODI | 0.11 | 0.13 | 0.12 | 0.23 | 1 |  |  |
| NDI | 0.05 | 0.06 | 0.07 | 0.29 | 0.31 | 1 |  |
| MFA | 0.23 | 0.26 | 0.54 | 0.68 | 0.66 | 0.66 | 1 |

RV coefficients, i.e., a multivariate extension of squared Pearson correlation coefficients. ADNI = Alzheimer's Disease Neuroimaging Initiative; fISO = free-water; MFA = multiple factor analysis; NDI = neurite density index; ODI = orientation dispersion index.

This can be further broken down by dimension, as is done in Supplementary Table 2, which provides the correlation of each parameter with the first and second dimensions and its coordinates and the relative contribution.

**Supplementary Table 2.**

|  | Correlation |  | Coordinates |  | Contribution |  |
| --- | --- | --- | --- | --- | --- | --- |
|  | Dim 1 | Dim 2 | Dim 1 | Dim 2 | Dim 1 | Dim 2 |
| Tau | 0.76 | 0.31 | 0.56 | 0.02 | 31.78 | 1.23 |
| fISO | 0.87 | 0.46 | 0.74 | 0.13 | 42.10 | 8.81 |
| ODI | 0.60 | 0.78 | 0.31 | 0.58 | 17.26 | 37.90 |
| NDI | 0.60 | 0.90 | 0.16 | 0.80 | 8.87 | 52.06 |

Additional information for the multiple factor analysis (MFA). In each case, each parameter's principal component analysis (PCA) results are related to the first and second dimensions (dim) of the MFA. Contribution refers to the percentage each variable contributes to the dimension in question and is thus a rough estimate of its importance. fISO = free-water; NDI = neurite density index; ODI = orientation dispersion index.

It is also helpful to recall that MFA performs a hierarchical process by decomposing subsidiary independent principal component analyses (PCAs). These analyses are presented in Supplementary Figure 2, where it can be seen that higher values of the first dimension of fISO and tau each predict significant declines in cognition. In comparison, higher ODI values are associated with increases in cognition (though these do not survive Bonferroni correction). Note that tau loads far more strongly than any other variable (i.e., its coordinates are all closer to one), and without the ability of MFA to weight or balance the variable sets, the result would be entirely dominated by this one variable. The second variable for each PCA is also presented (Supplemental Figure 2B). However, each explains far less variance and is generally not significant, save that higher values of fISO in frontal and striatal regions predict lower executive function values.

**Supplementary Table 3.**
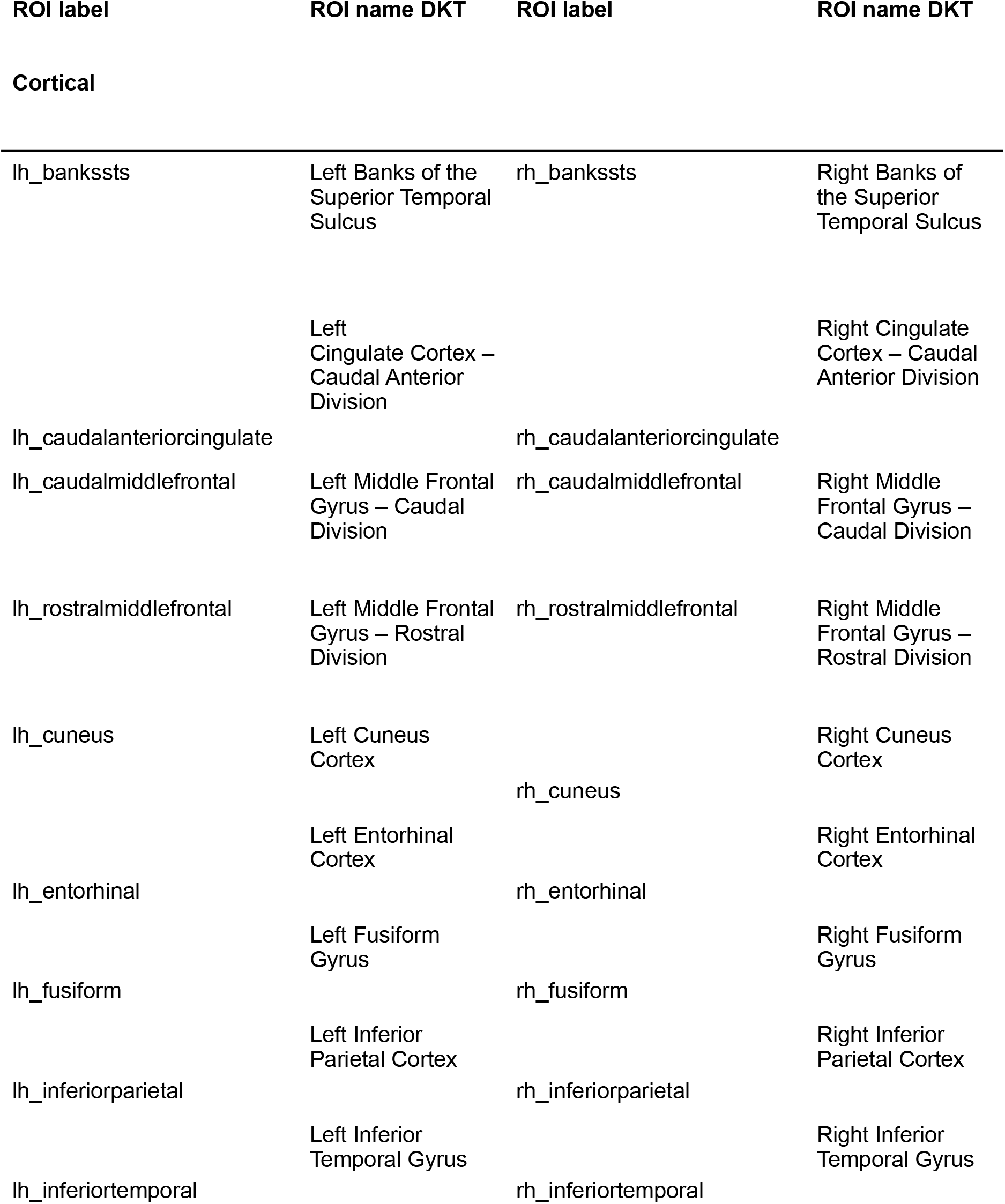
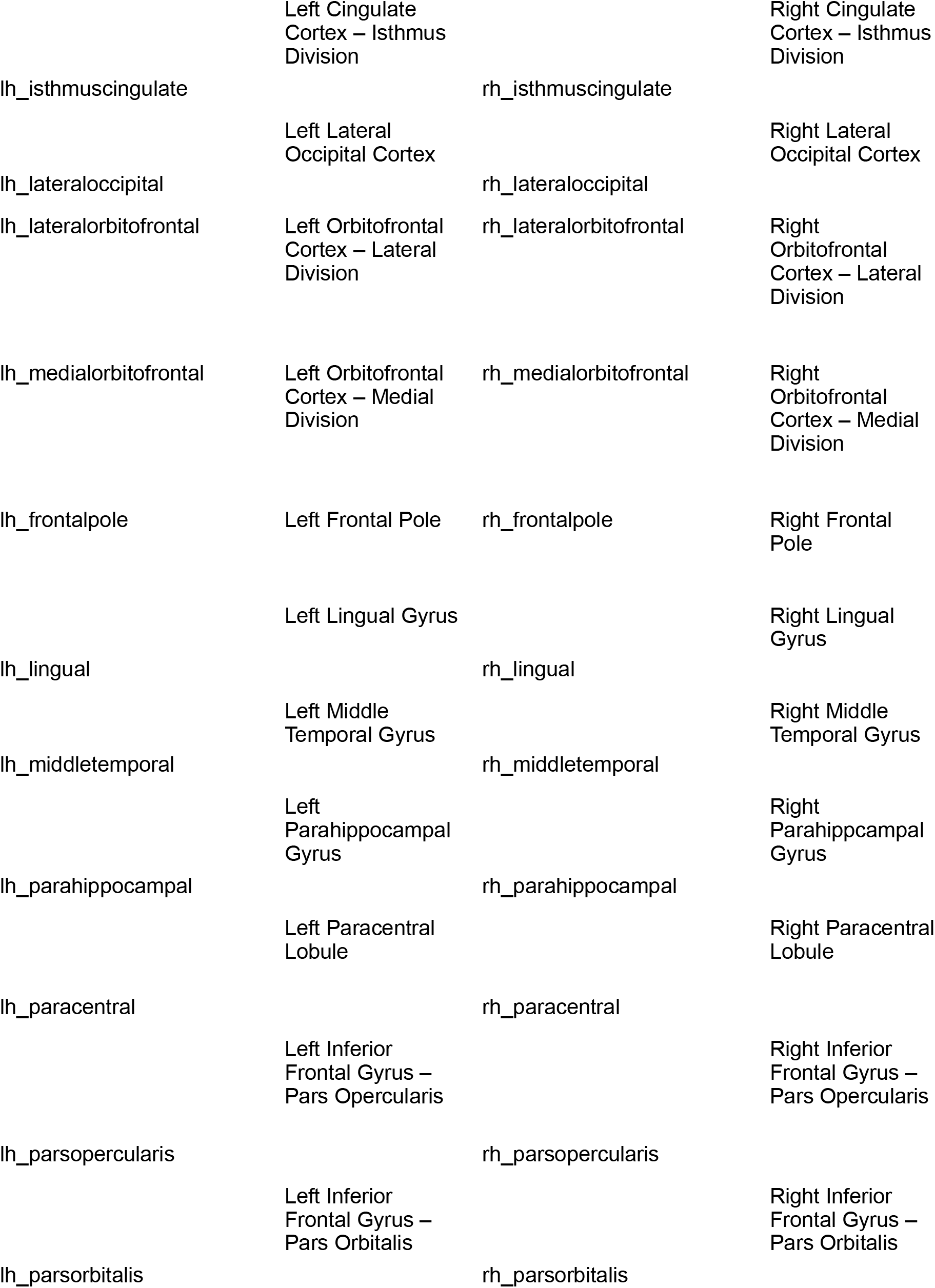
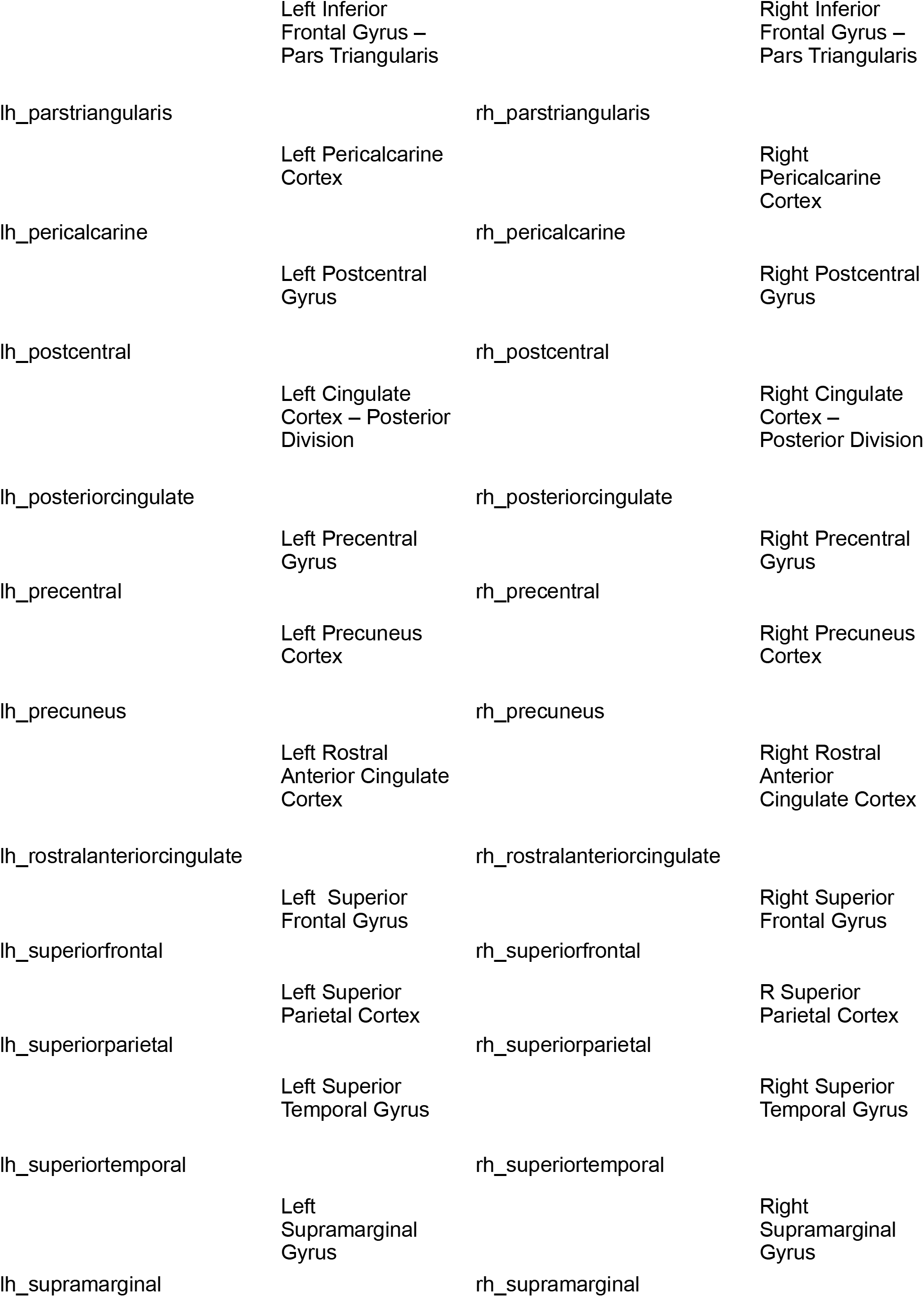
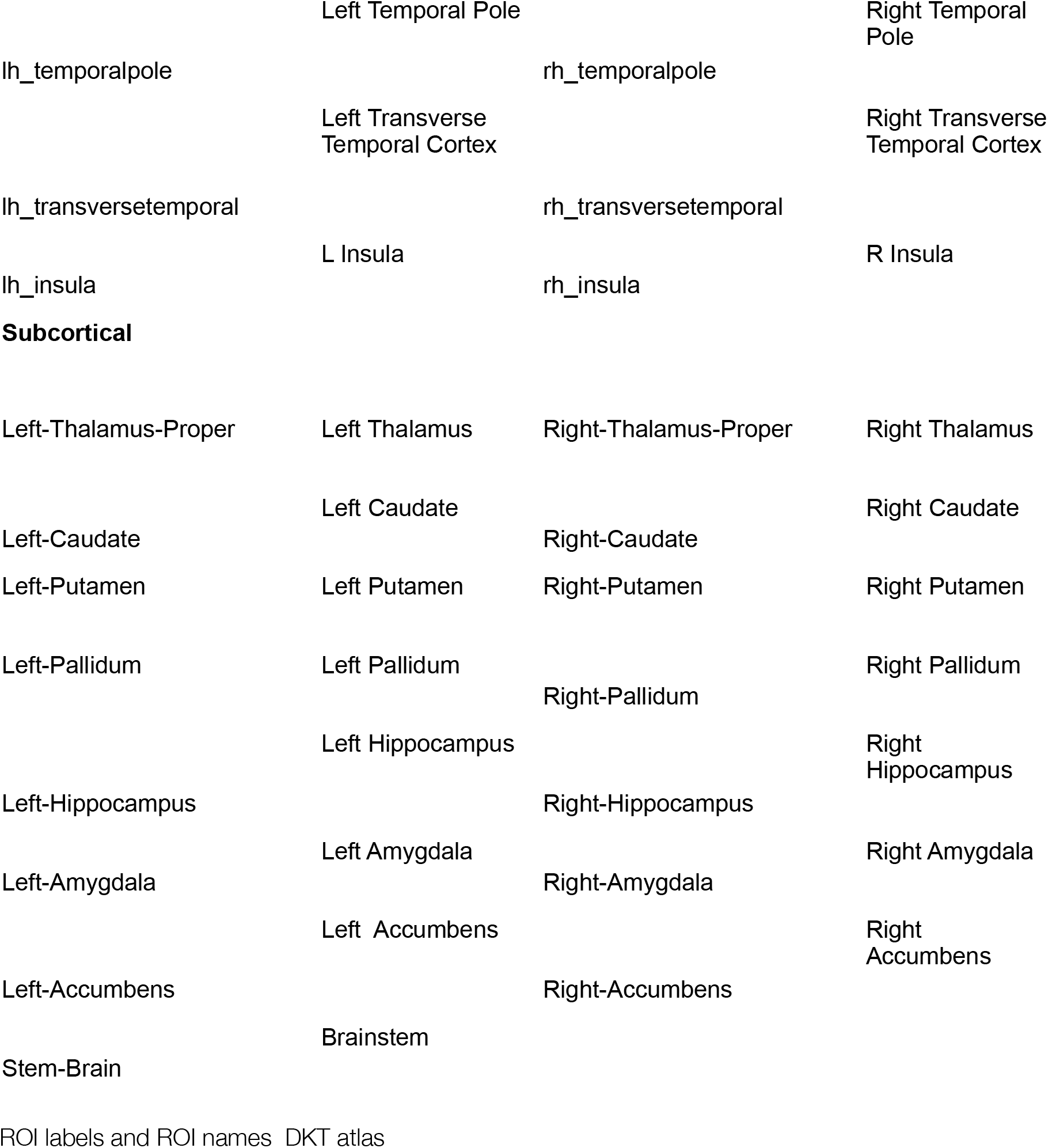

**Supplemental Figure 1.**
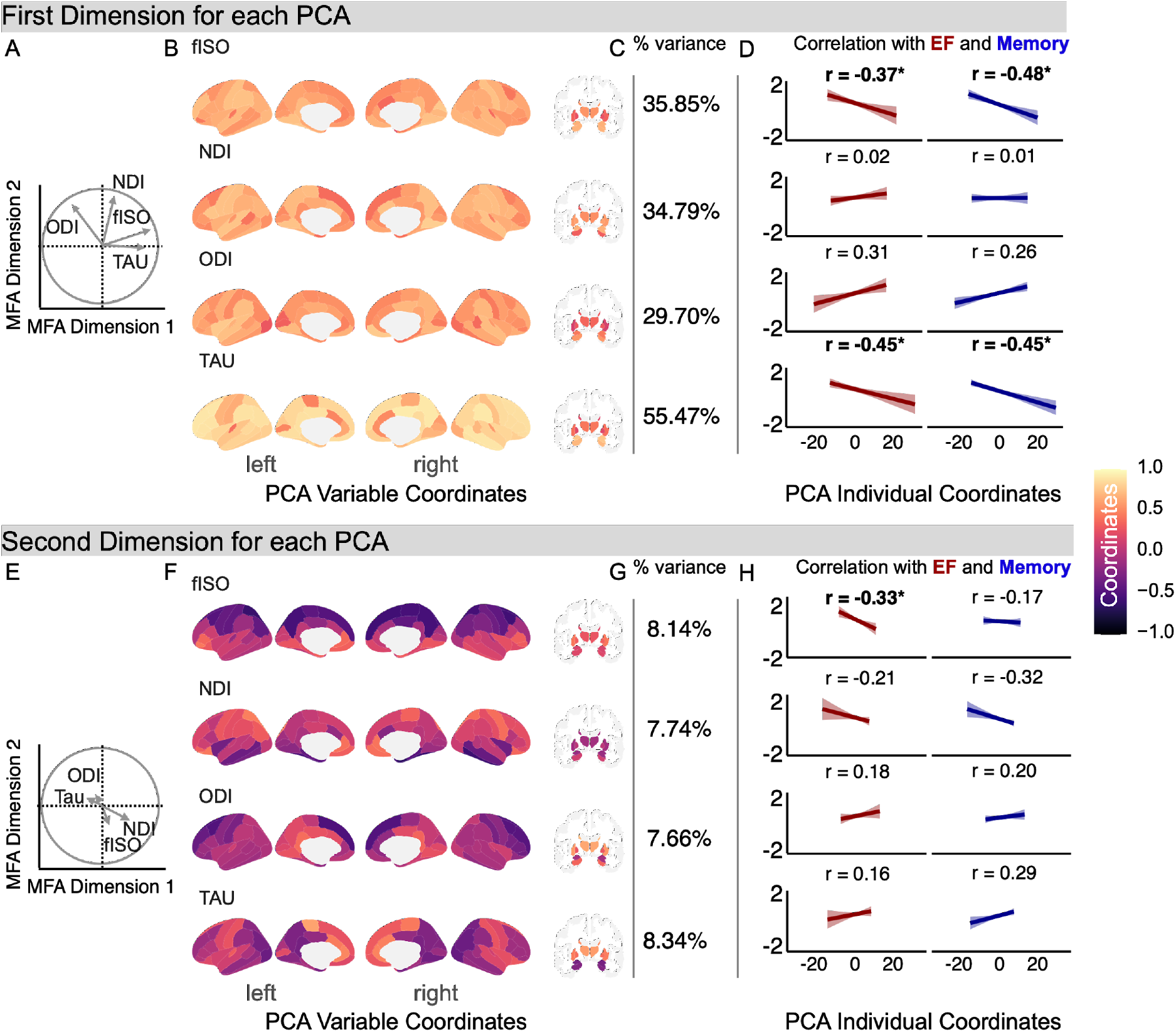
Breakdown of each principal component analysis (PCA) contributing to the global multiple factor analysis (MFA). The upper panel shows data from the first principal component (PC) for each parameter’s PCA, while the lower panel shows the second PC. Partial axes plots are shown for the first PCs A) and the second PCs B) in relation to the global MFA, answering how does each component from each modality-specific PCA relate to the global PCA. PCA coordinates are shown in brain-space in B) and F) and show regional variation in within-modality covariance, particularly on the second dimension. Each PCA’s percentage explained variance is shown in C) and G). Finally, subject-specific PC coordinate scores were used to predict behavior shown by the plots in D) and H). Asterisks and bolded lettering for correlations indicate significance FDR-corrected for 16 comparisons, at p = 0.05 level yielding a corrected p-threshold of 0.0031. EF = executive function; FDR = false discovery rate; fISO = free-water; NDI = neurite density index; ODI = orientation dispersion index

### Hierarchical Clustering on Principal Components (HCPC): Are there Clusters of Brain Parameters that Associate with Different Behavior Patterns?

MFA helps solve the multimodal data integration problem and reduces a complex set of parameters to a few dimensions. However, it is often useful to go one extra step and cluster on these dimensions. Thus we employ hierarchical clustering on the MFA output. We hoped participants might form data-driven clusters with relative neurite orientation dispersion and density imaging (NODDI) parameters and tau levels that vary in their relationship with cognition. We follow Husson et al. (2017) and employ v-tests(Husson *et al*., 2017), to test whether the average value of a quantitative variable within a category or cluster differed more than the general average (i.e., chance), and is expressed as a “standardized deviation between the mean of those individuals with[in] the category and the general average.” In this sense, a v-test can be interpreted similarly to a z-test and produces a significance test.

Examining a dendrogram suggested a three cluster-solution was optimal (see Supplemental Figure 3A), which agreed with the software’s suggestion. These clusters are also projected onto the first two dimensions of the MFA (Supplemental Figure 3B). Parameter values differing significantly by cluster, as determined with a v-test, were projected into brain space using the ggseg package (Supplemental Figure 3C). Cluster 1 is characterized by low values of all NODDI parameters and tau; cluster 2 is relatively undifferentiated in tau or fISO, but has higher NDI and ODI values than the other two clusters. Finally, the third cluster is characterized by higher fISO and tau values, with lower ODI values in the bilateral hippocampus, sensorimotor, medial prefrontal, and posterior cingulate cortices. For this third cluster, many of the same regions with lower ODI values also had higher NDI values.

We then used four general linear models to test whether cluster membership impacted the relationships between the first or second MFA dimensions with the ADNI factor scores (see Supplemental Figure 3D). For the first MFA dimension, the main effect of cluster membership was not significant (all *p*’s > 0.36) for either memory or executive function, nor were any interactions significant (all *p*’s > 0.15), though in each case, the main effects of the first dimension on memory *F* (1, 74) = 7.9, *p* =0.006, 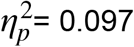, and executive function *F* (1, 74) = 5.6, *p* =0.02, 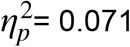, were significant suggesting that higher values of the first MFA dimension, representing higher fISO and tau, led to universally worse cognitive scores across participants.

**Supplemental Figure 2.**
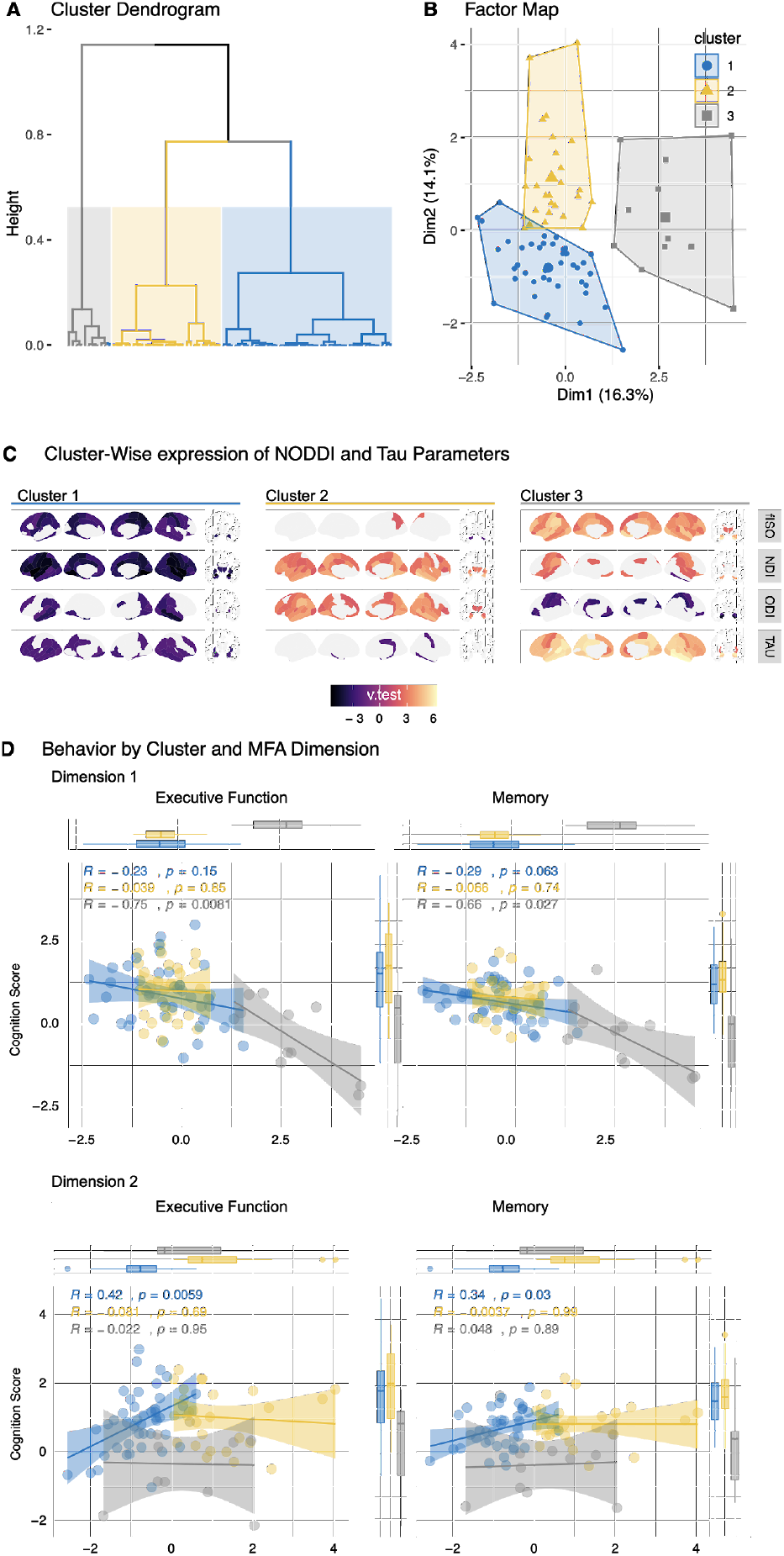
Hierarchical Clustering on Principal Components (HCPC). Clusters are plotted with a dendrogram A) to help justify a three-cluster solution and B) in the multiple factor analysis (MFA) component space for comparison with earlier results. Significant v-test statistics (similar to z-tests) are shown in brain space in C), and relationships between MFA components and cognition by the cluster is shown in D). Dim = dimension; fISO = free-water; NDI = neurite density index; ODI = orientation dispersion index.

For the second MFA dimension, the main effect of dimension (where higher values represented more NDI and ODI) was not a significant predictor of either memory or executive function (both *p*’s > 0.16). There were significant main effects of cluster membership in both the memory *F* (2, 74) = 15.01, *p* < 0.001, 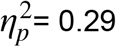 and executive function models *F* (2, 74) = 15.01, *p* < 0.001, 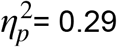, driven in each case by lower cognitive scores in cluster three. Post-hoc tests showed that both the first and second cluster had significantly higher memory and executive function scores than the third cluster (all Bonferroni-corrected *p*’s < 0.001). There was also a significant interaction between cluster membership and second dimension scores on executive function, *F* (2, 74) = 3.18, *p*=0.047, 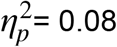, such that in the first cluster, higher values of the second MFA dimension (representing higher values of NDI and ODI) corresponded to better executive function performance (see Figure 2D).

## Footnotes

1 Please note that wherever cortical thickness is referred to, this also includes subcortical volume.

